# Receiving information on machine learning-based clinical decision support systems in psychiatric services may increase patient trust in these systems: A randomised survey experiment

**DOI:** 10.1101/2024.04.11.24305655

**Authors:** Erik Perfalk, Martin Bernstorff, Andreas Aalkjær Danielsen, Søren Dinesen Østergaard

**Affiliations:** Department of Affective Disorders, Aarhus University Hospital – Psychiatry, Aarhus, Denmark; Department of Clinical Medicine, Aarhus University, Aarhus, Denmark

## Abstract

**Background:** Clinical decision support systems based on machine learning (ML) models are emerging within psychiatry. If patients do not trust this technology, its implementation may disrupt the patient-clinician relationship. Therefore, we examined whether receiving basic information about ML-based clinical decision support systems increased trust in them.

**Methods:** We conducted an online randomised survey experiment among patients receiving treatment in the Psychiatric Services of the Central Denmark Region. The participants were randomised to one of three arms, receiving different types of information: Intervention = information on clinical decision making supported by an ML model; Active control = information on a standard clinical decision process without ML-support; Blank control = no information. The participants were unaware of the randomization and the experiment. Subsequently, the participants were asked about different aspects of trust/distrust in ML-based clinical decision support systems. The effect of the intervention was assessed by comparing pairwise comparisons between all arms on component scores of trust and distrust.

**Findings:** Out of 5800 invitees, 992 completed the survey experiment. The intervention increased trust in ML-based clinical decision support systems when compared to the active control (mean absolute difference in trust: 5% [95%CI: 1%;9%], p-value= 0·009) and the blank control arm (mean absolute difference in trust: 4% [1%;8%], p-value=0·015). Similarly, the intervention significantly reduced distrust in ML-based clinical decision support systems when compared to the active control (mean absolute difference in distrust -3%[-5%; -1%], p-value=0·021) and the blank control arm (mean absolute difference in distrust -4% [-8%; -1%], p-value=0·022). For both trust and distrust, there were no material or statistically significant differences between the active and the blank control arms.

**Interpretation:** Receiving information on ML-based clinical decision support systems in hospital psychiatry may increase patient trust in such systems. Hence, implementation of this technology could ideally be accompanied by information to patients.

**Funding:** None.

**Research in context:** *Evidence before this study:* Clinical decision support systems based on machine learning (ML) models are emerging within psychiatry. However, if patients do not trust this technology, its implementation may disrupt the patient-clinician relationship. Unfortunately, there is only little knowledge on opinions on ML models as decision support among patients receiving treatment in psychiatric services. Also, it remains unknown whether receiving basic information about ML-based clinical decision support systems increases patients’ trust in them. We searched PubMed on Sep 12, 2023, with the terms “((survey) OR (survey experiment)) AND (patients) AND ((opinions) OR (attitudes) OR (trust)) AND ((machine learning) OR (artificial intelligence)) AND ((Psychiatry) OR (Mental Disorders) OR (Mental Health))” with no language restrictions. This yielded a total of 73 records, none of which surveyed a patient population from psychiatric services. Only two studies were directly relevant for the topic at hand. One surveyed patients from a general hospital system in the United States about the use of ML-based prediction of suicide risk based on electronic health record data. The results showed that patients were generally supportive of this data use if based on consent and if there was an opportunity to opt out. The other study surveyed women from the general population about their opinion on the use of artificial intelligence (AI)-based technologies in mental healthcare. The results showed that the respondents were generally open towards such technologies but concerned about potential (medical harm) and inappropriate data sharing. Furthermore, the respondents identified explainability, i.e., understanding which information drives AI predictions, as being of particular importance.

*Added value of this study:* To the best of our knowledge, this is the first study to investigate opinions on ML-based clinical decision-support systems among patients receiving treatment in psychiatric services. On average, patients were open towards the use of ML-based clinical decision-support systems in psychiatry. Furthermore, the results suggest that providing basic information about this technology seems to increase patient trust in it, albeit with a small effect size. Finally, the results support prior reports on the importance of explainability for acceptance.

*Implications of all the available evidence:* Receiving information on ML-based clinical decision support systems in hospital psychiatry, including how they work (explainability), may increase patient trust in such systems. Hence, successful implementation of this technology likely requires information of patients.

## Introduction

Machine learning (ML) is based on the idea that machines (computers) can learn from historical data and be trained for pattern recognition (e.g., prediction). The perspectives of using ML to aid decision-making in the medical field are promising.^1^ Indeed, prediction models based on ML have shown to be accurate in many clinical contexts with performance levels comparable to- or above those of clinicians.^2^

Since patients are major stakeholders in the medical field, their acceptance of ML is of paramount importance with regard to successful implementation of tools based on ML.^3,4^ Public and patient trust in ML in the healthcare setting has been surveyed before.^6–8^ According to these studies, stakeholder trust in medical application of ML models relies on the knowledge that the final decision lies in the hands of the health professionals, i.e. that ML models are merely used for decision *support*. Moreover, increased information about ML models, including explainability (transparency of what drives prediction), is associated with increased trust in them.^7,9,10^

To our knowledge, however, no prior surveys have focused on opinions on ML-based clinical decision support systems among patients receiving treatment in psychiatric services. This is an unfortunate gap in the literature, as such systems are gaining traction in the psychiatric field^11,12^ and because the level of general and institutional trust is relatively low among some patient groups in psychiatry.^13,14^ Therefore, the aim of the present study is to investigate trust in ML-based clinical decision support systems among patients with mental disorders. Furthermore, we will test whether receiving information about ML-based clinical decision support systems will increase patients’ trust in this technology.

## Methods

### Design

We conducted a randomized online survey experiment focusing on ML-based clinical decision support tools in psychiatric services. The study design and analysis plan were pre-registered and are available at https://osf.io/y5n3a/?view_only=a5c42a476eee4cd5996b35963b00ae60. The design of the study is illustrated in Figure 1.

**Figure 1.**
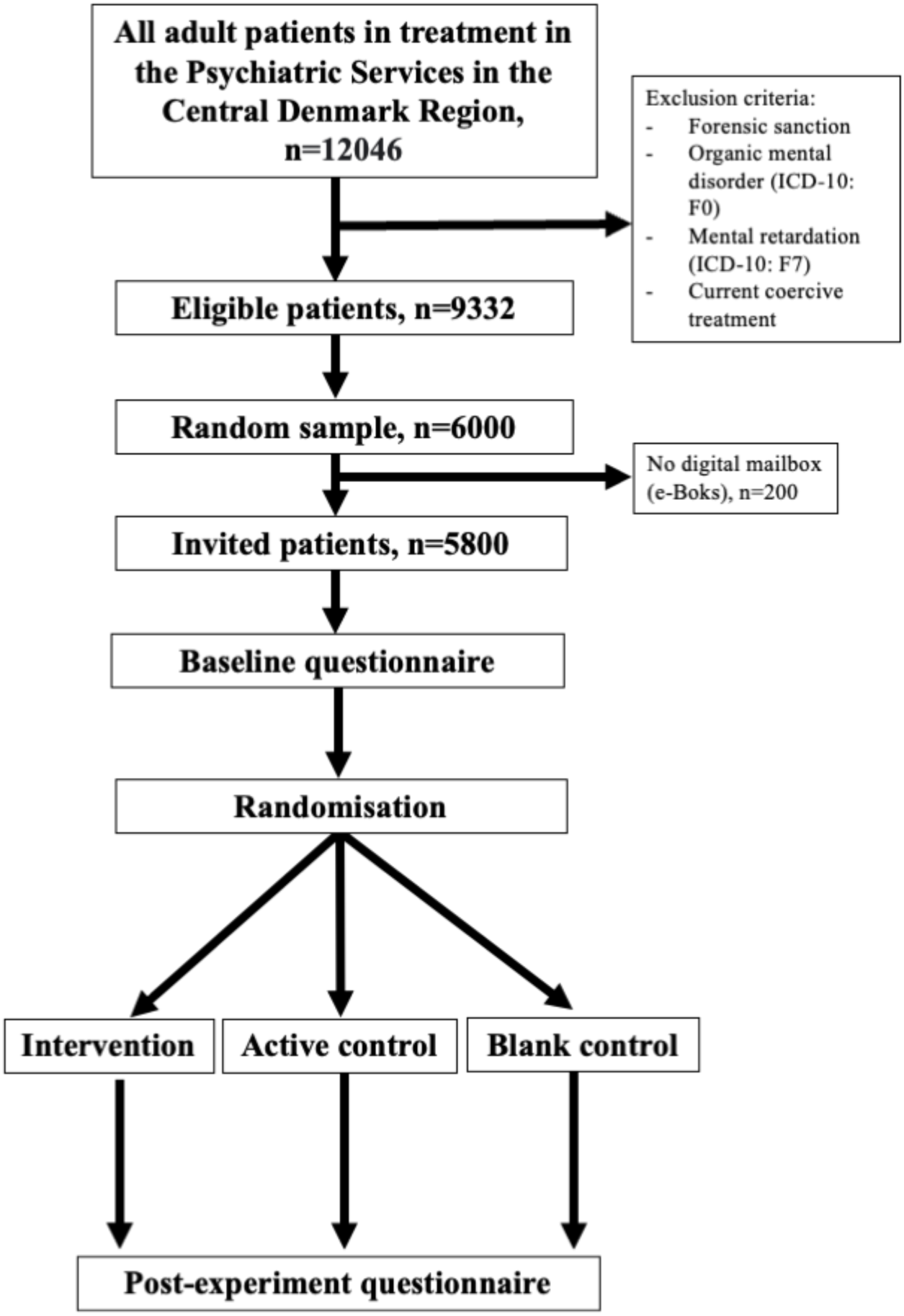
Flowchart of study design and population e-Boks: The secure digital mailing system used by Danish authorities to communicate with citizens

### Setting

The survey experiment was performed within the Psychiatric Services of the Central Denmark Region, which has a catchment area of approximately 1·3 million people. It comprises five public psychiatric hospitals, which provide free (tax-financed) inpatient, outpatient, and emergency psychiatric treatment.

### Participants

Patients were eligible for participation if they were 18 years old and received treatment in the Psychiatric Services of the Central Denmark Region. Patients were ineligible if they had a forensic sanction, received coercive treatment, or had an organic mental disorder (ICD-10 code: F0X.X), or mental retardation (ICD-10 code: F7X.X). Based on power calculations (see below), we invited 6000 randomly drawn eligible patients using the online survey service SurveyXact^15^) via “eBoks“ – the secure digital mailing system used by Danish authorities to communicate with citizens.^16^ The survey was distributed from May 26-31, 2023. A reminder was sent on June 12, 2023, to those not having responded yet. The participants provided informed consent for study participation by ticking a box and entering their unique social security number and name. The participants did not receive any monetary incentive for participation.

After the survey was fielded, it came to our attention that participants using an Android-based device to access the “e-Boks” app could not access the hyperlink provided in the invitation. To solve this issue, we informed the participants of a solution to this technical problem when distributing the reminder. While this issue is likely to have reduced the response rate, we consider it unlikely to have introduced bias.

### Power calculation

A power-analysis (alpha= 0·05, power= 0·80, two-sided) for a pair-wise comparison, assuming a small intervention effect of Cohen’s D = 0·2 (as seen in similar studies^9^), estimated that 1200 participants (400 per randomization arm) were required. A recent study in the same setting and using the same invitation procedure had a response rate of approximately 20%.^17^ Based on this effect size and response rate, 6000 patients were invited to participate in the study.

### Randomisation

Participants were randomly allocated to one of three arms: intervention, active control, and blank control (for a description, see the “Survey experiment” section, below). As SurveyXact did not allow for standard 1/3, 1/3, 1/3 random allocation at the time of the study, the system was devised to assign the participants to one of the three arms based on the time they accessed the survey link in the invitation letter. Specifically, participants accessing the link in the intervals from >=0 to 0.333 second; from >0.333 to 0.666 second, and from >0.666 to <1 second were assigned to the blank control arm, the active control arm, and the intervention arm, respectively. The participants were not aware that a randomisation and intervention took place.

### Baseline questionnaire

When entering the survey, irrespective of allocation arm, the participants initially filled in a baseline questionnaire regarding education level, current work status, household composition (adults and children), general trust, trust in technology, and perceived understanding of standard clinical decision making as well as perceived understanding of ML-based clinical decision support systems. Answers on trust and perceived understanding were provided on Likert scales from 0-10 (an English translation of the questionnaire in Danish is available in Supplementary Table 1).

### Survey experiment

The baseline questionnaire was followed by the experiment in which the participants received three different types of information based on the randomised allocation:

1. Intervention: Visual and text-based information pamphlet (slides within the electronic survey – see Supplementary Table 2) explaining how an ML-based clinical decision support system works and may aid clinical practice in psychiatric services.
2. Active control: Visual and text-based information pamphlet (slides within the electronic survey – see Supplementary Table 3) explaining a standard clinical decision process in psychiatric services without the use of a ML-based clinical decision support system.
3. Blank Control: No information pamphlet.

### Post-experiment questionnaire (outcome measure)

After the survey experiment, the participants filled in a questionnaire aimed at measuring trust and distrust in ML-based clinical decision support systems in psychiatric services. Specifically, the respondents answered the following questions: 1. “I feel safe that mental health professionals can make decisions with the support of machine learning models.” 2. “I trust that the Psychiatric Services can use machine learning models in a safe and appropriate way.” 3. “I am concerned that use of machine learning models for decision support in psychiatry will increase the risk of error.” 4.” I would like to have the opportunity to opt out of machine learning models being used for decision support in relation to my treatment in the psychiatric services.” 5. “I am concerned that healthcare services, including the psychiatric services, are becoming too dependent on machine learning models.” 6. “I am concerned that the use of machine learning models may lead to increased inequality in healthcare, including psychiatry.” 7. “The advantages of using machine learning models for decision support in psychiatry outweigh the disadvantages.” 8. “It is important to me that I can get an explanation of the basis on which a machine learning model recommends a given treatment”. 9. ”I am concerned that a machine learning model may make incorrect recommendations due to inaccuracies in my medical record.” The questions were adapted from prior studies covering the same topic.^7,18^All questions were answered using an 11-level Likert scale ranging from 0 (“Totally disagree”) to 10 (“Totally agree”).

### Choice of primary outcome measure

As it is suboptimal to sum positively- and negatively-worded items (after inversion),^19^ the three positively-worded “trust” items (1, 2, and 7)) and the 5 negatively-worded “distrust” items (3, 4, 5, 6, and 9) were grouped a priori (see the pre-registered analysis plan). Item 8 was considered to be neutral and was therefore kept separate. Subsequently, principal component analyses were performed to test whether the trust and distrust items, respectively, loaded onto latent components. The number of components was determined by analyzing the scree plot and choosing the number of components before the distinct break (“elbow”) in the plot.^20^ Subsequently, an item was considered to load onto a component if it had a loading of >0.40 or <-0.40.^21^ Based on the number of items loading onto the component, a trust total score (the sum of the positively-worded items) and a distrust total score (the sum of the negatively-worded items) were constructed to be used as the outcome measures.

### Handling of survey responses

After clicking the generic link to the survey, the patients identified themselves by manually inserting their social security number and their name. If the participants completed the questionnaire more than once, the first response was used. If the participants first made a partial response, including the randomization element, and subsequently completed the survey, while randomized to a different arm, then the full response was not included in the analyses (as these participants were unblinded to the randomization). As the outcome measures was placed at the end of the survey, only completers were included in the analyses. Questions could not be left blank. Thus, there were no missing values.

### Supplementary data from electronic health records

The participants gave consent to extraction of sociodemographic (age, sex, civil status) and clinical data (the number of contacts to the psychiatric services including contact type (inpatient/outpatient) and the associated ICD-10 diagnoses, as well as the time since their first contact to the psychiatric services) from the electronic health records for the purpose of the study. Linkage was performed using the respondent’s unique personal identification number, ^22^ To define diagnostic subgroups, we considered the participant’s most severe main diagnosis (registered from 2011 until the time of the survey) using the following ICD-10 hierarchy: F2x (psychotic disorders) > F3x (mood disorders) > F4x (anxiety- and stress-related disorders) > F5x (eating, sleeping, and other behavioural syndromes associated with physiological disturbances) > F6x (personality disorders) > F8x (developmental disorders including autism) > F9x (child and adolescent mental disorders) > F1x (substance use disorders).

### Statistics

The sociodemographic and clinical characteristics were summarized using descriptive statistics. Potential differences in time used for survey completion between the randomization groups was tested using Mann-Whitney U test. As primary analyses, the level of trust/distrust in ML-based clinical decision support systems quantified by the trust/distrust scores were compared pairwise between the three randomization arms via two-sample t-tests. Equivalent, secondary analyses were conducted at the individual trust/distrust item level. The Pearson correlation coefficient was used to assess correlation between the latent trust and distrust mean scores. As robustness analyses, linear regression analysis of trust/distrust in ML-based clinical decision support systems was performed across the three randomization arms, while stratifying by sex, age, diagnostic groups, socioeconomic factors (e.g., educational level, current work status), baseline knowledge of machine learning as decision support, and the level of general trust. The significance threshold was set at 0·05. Correction for multiple comparisons was not performed as the analyses were pre-registered and highly interdependent.^23^ All data management and statistical analyses were performed using Rstudio version 2023·06·0 Build 421.

### Ethics

Research studies based on surveys are exempt from ethical review board approval in Denmark (waiver no. 1-10-72-138-22 from the Central Denmark Region Committee on Health Research Ethics). The study was approved by the Legal Office in the Central Denmark Region (reg. no. 1-45-70-21-23) and registered on the internal list of research projects having the Central Denmark Region as data steward (reg. no. 1-16-02-170-23). Prior to the survey, to ensure that it was appropriate for the study population, we received feedback on the questionnaire and the “intervention” and “active control” information pamphlets from two patients having received treatment in the Psychiatric Services in the Central Denmark Region.

## Results

A total of 992 invitees completed the survey (106 partial respondents were excluded) and Table 1 lists their sociodemographic and clinical characteristics as well as baseline information regarding trust in institutions and technologies. See Supplementary Table 4 for clinical characteristics of 1098 randomized participants (Full respondents + partial respondents).

**Table 1:**
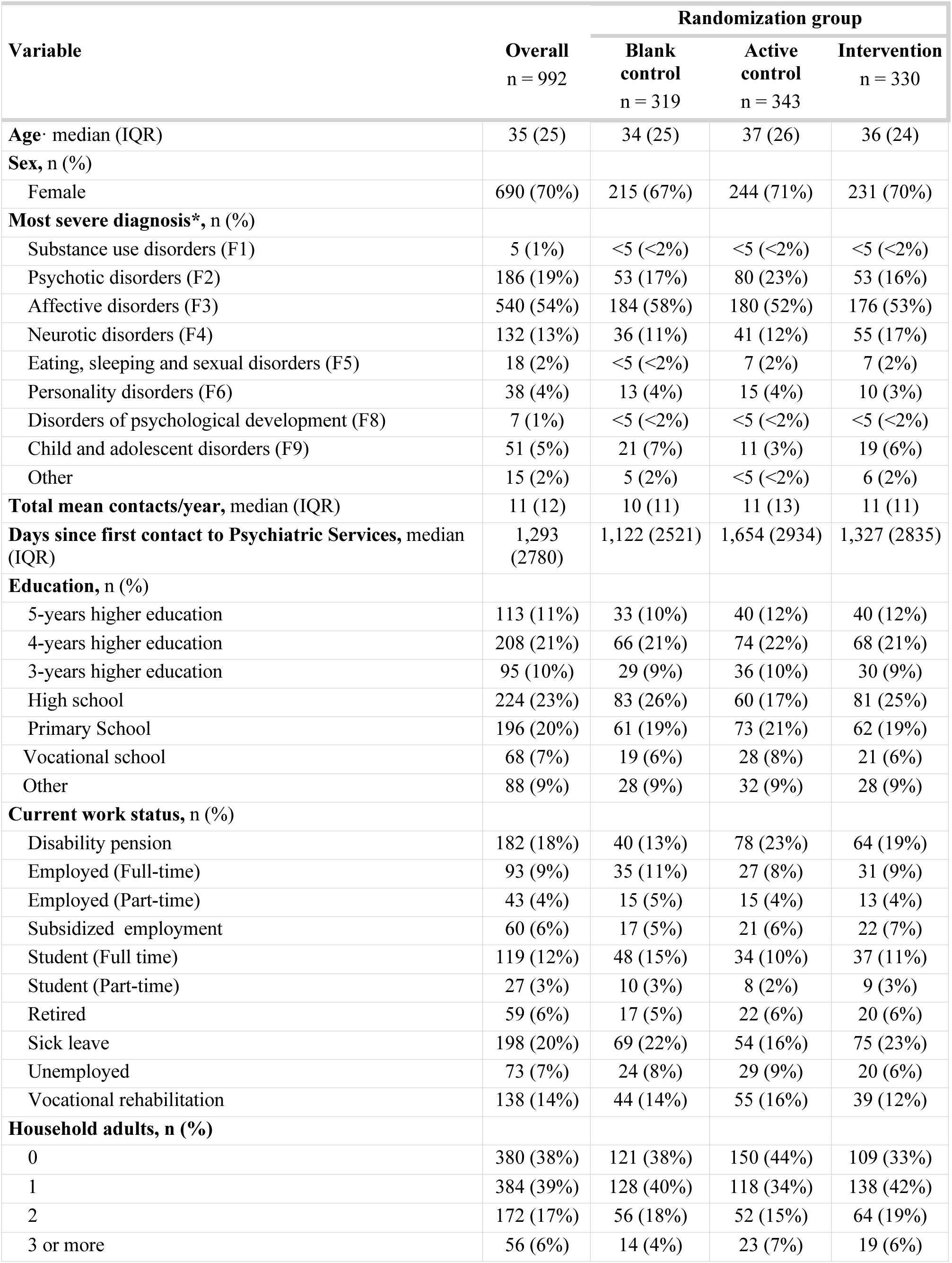
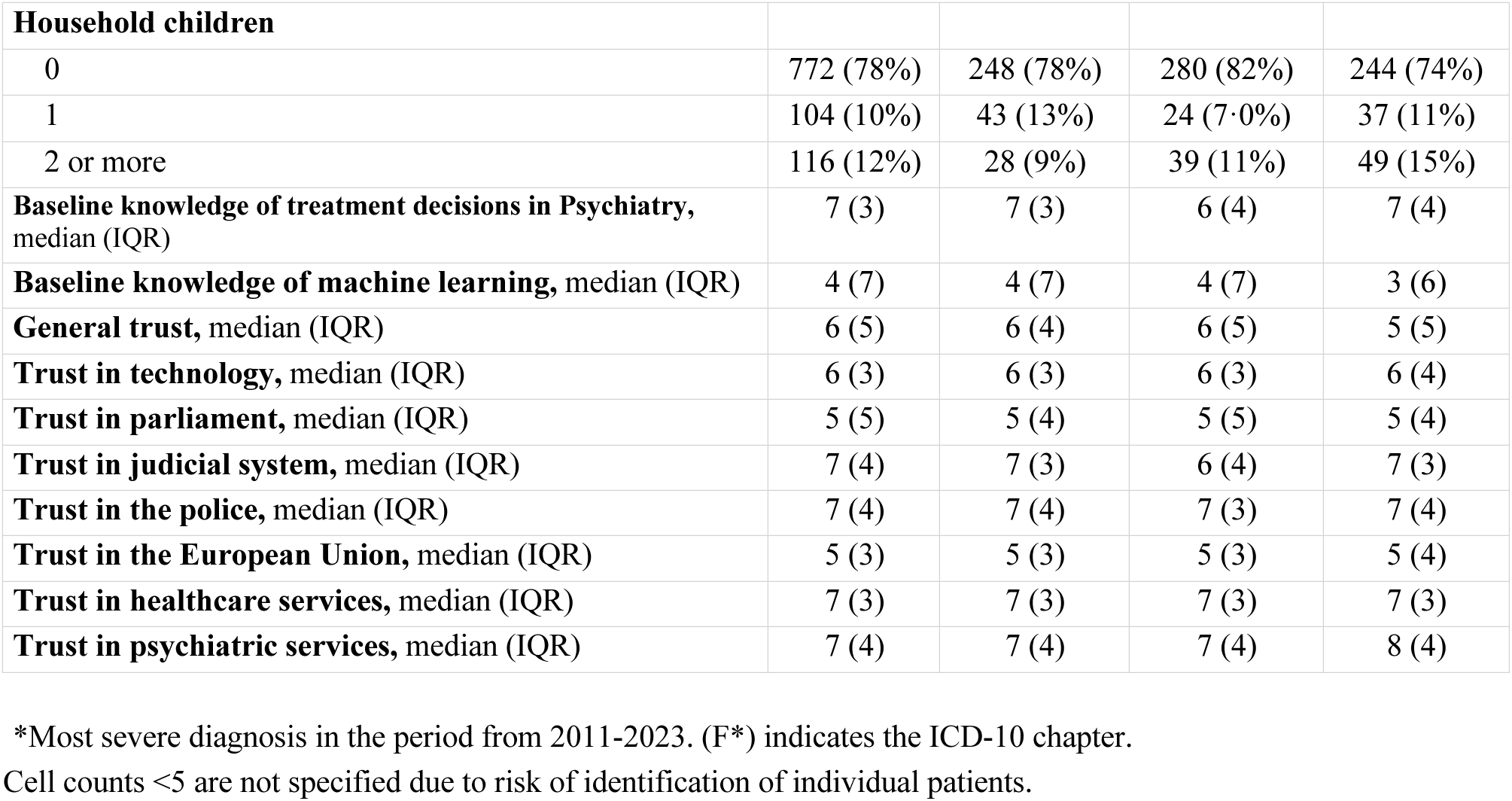
Characteristics of the 992 participants with complete responses.

The randomization led to the following distribution of full respondents across arms: blank control=319 (partial respondents=37), active control=343 (partial respondents=33) and intervention=330 (partial respondents=36). The median response time was 280 seconds (IQR: 167) for those allocated to the blank control arm, 368 seconds (IQR: 264) for the active control arm, and 388 seconds (IQR: 244) for the intervention arm. The response time for those in the active control arm and the intervention arm were statistically significantly longer than for those in the blank control arm (p-values of <0·0001 on both tests) but did not differ statistically significantly from each other (p-value=0·18).

The results of the principal component analysis are shown in Supplementary Tables 5-6 and Supplementary Figures 1-2. As expected a priori, the survey items were grouped into a trust component consisting of the three positively worded items and a distrust component consisting of the five negatively worded items). The trust and distrust sum scores were inversely correlated (Pearson correlation coefficient= -0·60).

Table 2 shows the response to the items focusing on trust and distrust in ML-based clinical decision support systems, across the three randomization groups. Notably, the median scores on the trust items were generally within range of the trust general/institutional trust levels reported at baseline (available in Table 1).

**Table 2:** Individual item scores after the experiment.

| **Outcome items**<br>**(abbreviated\*)** | **Blank control** | **Active control** | **Intervention** |
| --- | --- | --- | --- |
| **Trust items** |  |  |  |
| Item 1: I feel safe with ML models, *median (IQR)* | 6 (4) | 6 (4) | 6 (5) |
| Item 2: Trust in ML models, *median (IQR)* | 6 (4) | 5 (4) | 7 (4) |
| Item 7: The advantages outweigh disadvantages, *median (IQR)* | 5 (3) | 5 (3) | 5 (3) |
| **Distrust items** |  |  |  |
| Item 3: Malpractice due to ML, *median (IQR)* | 5 (4) | 6 (5) | 5 (5) |
| Item 4: Possibility to opt out of ML, *median (IQR)* | 7 (5) | 6 (5) | 5 (6) |
| Item 5: Dependency on ML, *median (IQR)* | 6 (5) | 7 (5) | 6 (4) |
| Item 6: Inequality in healthcare due to ML, *median (IQR)* | 5 (4) | 5 (5) | 5 (5) |
| Item 9: Errors in EHR, *median (IQR)* | 8 (5) | 8 (5) | 8 (5) |
| **Neutral item** |  |  |  |
| Item 8: Explainability of ML models, *median (IQR)* | 10 (2) | 10 (2) | 10 (2) |

ML: Machine learning. EHR: Electronic health record. Mean differences between groups for single items and results from T-tests are shown in Supplementary Table 8. \*The full phrasing of the items and their scoring range are available in the methods section.

Figure 2 shows the results of the primary analyses, which compares the three randomization groups with regard to the trust and the distrust sum score. The intervention increased trust in ML-based clinical decision support systems when compared to the active control (mean absolute difference in trust: 5% [95%CI: 1%;9%], p-value= 0·0096) and the blank control arm (mean absolute difference in trust: 4% [1%;8%], p-value=0·015). Similarly, the intervention reduced distrust in ML-based clinical decision support systems when compared to the active control (mean absolute difference in distrust -3% [-5%; -1%], p-value=0·021) and the blank control arm (mean absolute difference in distrust -4% [-8%; -1%], p-value=0·022). For both trust and distrust, there were no material or statistically significant differences between the active and the blank control arms. The equivalent results at the level of the Individual trust and distrust items are listed in Supplementary Table 7 and are agreement with those from the analyses at the trust and distrust sum scores. Notably, the neutral item (importance of explainability of ML models) had an overall median of 10 (8-10) with a statistically significant difference between the blank control and the intervention arms (higher in the intervention arm), but not for the other comparisons.

**Figure 2.**
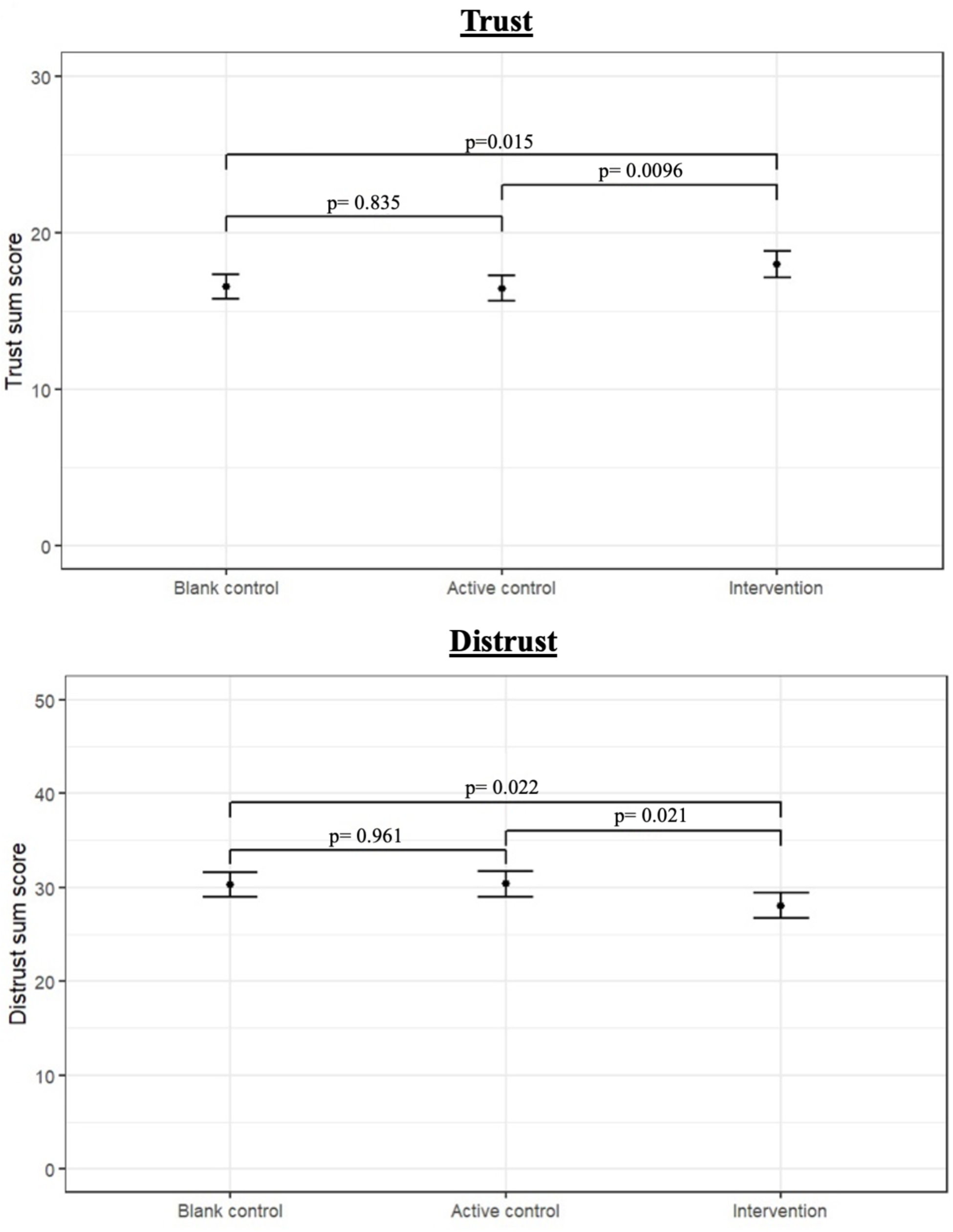
Effect of the intervention on trust (top) and distrust (bottom) in machine learning model-based clinical decision support systems

The results of the primary analyses stratified by sex, age, diagnostic groups, educational level, current work status, baseline knowledge of machine learning as decision support, and the level of general trust are listed in Supplementary Table 8 and suggest that the intervention effect is consistent across these strata.

## Discussion

This randomized survey experiment among patients receiving treatment in psychiatric services showed that information on ML-based clinical decision support systems may increase patient trust in such systems. Notably, the results regarding the explainability of ML models suggest that this aspect was particularly important for the respondents.

To the best of our knowledge, this study represents the first investigation into whether providing information about ML as a decision-support tool may enhance patient trust in such systems, suggesting that this is indeed the case. This result is in line with that from the study by Cadario et al., predominantly targeting individuals from the general population, which investigated the effect of receiving information on the variables (shape/size/colour) driving a malignant melanoma risk prediction algorithm.^9^ In the study by Cadario et al, the participants either received information on human healthcare providers versus ML algorithm’s decision-making processes. Consistent with our study, it was found that a small amount of information about ML algorithms reduced “algorithm aversion”, i.e., reduced the reluctance to utilize algorithmic support compared to human providers.^9^ The results from Cadario et al. also suggest that explainability is positively associated with uptake of prediction algorithms at the general population level. This is consistent with the patient perspective observed in the present study in which the respondents agreed the most with the item stating “It is important to me that I can get an explanation of the basis on which a machine learning model recommends a given treatment.” Analogue findings have been reported in studies of patients (within, e.g., primary care, radiology and dermatology)^6^, clinicians (within, e.g. internal medicine, anaesthesia, and psychiatry)^24^, and the general population.^6,7^ Taken together, these findings suggest that when developing ML models for healthcare, there should be emphasis on explainability to ensure trust among both clinicians and patients.

While the results of the present study are indicative of a causal effect of the information intervention, the effect was numerically quite small. This was expected as the intervention consisted of an electronic pamphlet with only four slides of text and pictures, which was administered only once. Furthermore, it can also be argued that the effect of the intervention may well be short-lived as the trust/distrust outcome was measured immediately after the intervention. For these reasons, future studies should ideally employ more comprehensive and repeated interventions, and longer time between the interventions and the outcome measurement.

Another aspect potentially affecting the effect size of the intervention is the timing of the survey. Specifically, as the survey was fielded at the end of May 2023, it came in the aftermath of the press coverage of the open letter signed by several high-profile tech leaders calling for a pause in the development of artificial intelligence until appropriate safeguards and legislation would be in place.^25^ This media coverage primarily focused on the security concerns and potential hazards associated with this emerging technology.^26,27^ This somewhat negative press on artificial intelligence could have rendered the respondents resistant to the information in the intervention (reduced effect size). In contrast, however, it is also possible that the negative press had resulted in reduced baseline trust in the technology, which would then leave ample room for a positive effect of the information conveyed in the intervention (increased effect size). Based on the data at hand, we are, unfortunately, not able to determine the overall direction of this potential response bias.

There are limitations to this study, which should be taken into account by the readers when interpreting the results. First and foremost, the survey had a relatively low response rate, which means that selection bias may be in play. Due to the lack of clinical and sociodemographic data on those not participating, we were unable to adjust the analyses for attrition. However, in a recent 2-wave survey in the same population during the COVID-19 pandemic (focusing on psychological distress/well-being among patients with mental disorders during the pandemic), we were granted force majeure access to clinical and sociodemographic data on non-respondents. This allowed us to employed inverse probability weighting to address the potential bias arising from non-response. Notably, this adjustment had no material impact on the results.^17,28^ While this does not preclude selection bias in the present study (a different survey topic and timing), it does suggest that this is unlikely to be a substantial problem. Second, we did not use a validated questionnaire for measuring trust and distrust in ML as a clinical decision support tool as such questionnaires, to our knowledge, have not yet been developed. With this inherent limitation in mind, we believe that the latent components of trust and distrust derived from principal component analysis of items with apparent face validity, represent reasonable outcome measures. Third, we did not have attention check questions, which means that some participants may have responded inconsistently/arbitrarily to the questions. This would have introduced noise in the data and complicated signal detection. Yet, a signal (the intervention increasing trust and reducing distrust) was indeed detected. Fourth, with regard to generalizability, it should be borne in mind than Denmark is among the most digitalized countries in the world,^29^ and its inhabitants may, therefore, be more positive towards new technology. Thus, replication of the reported findings in other countries is warranted.

In conclusion, this survey experiment suggests that receiving information on ML-based clinical decision support systems in hospital psychiatry likely increases patient trust in such systems. This is compatible with results from studies of other patient populations, clinicians as well as the general population. Thus, when taken together, the literature suggests that providing appropriate information to patients will be of importance when implementing ML-based clinical decision support systems.

## Contributors

All authors contributed to the conceptualization and design of the study. EP and SDØ conducted and supervised the data collection. Data management, verification, and statistical analysis was carried out by EP. All authors contributed to the interpretation of the obtained results. EP wrote the first draft of the manuscript, which was subsequently revised for important intellectual content by the other authors. All authors approved the final version of the manuscript prior to submission.

## Data Availability

The data cannot be shared as the participants have not consented to data sharing.

## Acknowledgments

There was no specific funding for this study. Outside this study, SDØ reports funding from the Lundbeck Foundation (grants R358-2020-2341 and R344-2020-1073), the Novo Nordisk Foundation (grant NNF20SA0062874), the Danish Cancer Society (grant R283-A16461), the Central Denmark Region Fund for Strengthening of Health Science (grant 1-36-72-4-20), the Danish Agency for Digitisation Investment Fund for New Technologies (grant 2020-6720), and Independent Research Fund Denmark (grant 7016-00048B and 2096-00055A). These funders played no role in the design or conduct of the study; collection, management, analysis, and interpretation of the data; preparation, review, or approval of the manuscript; and decision to submit the manuscript for publication.

The authors are grateful to Bettina Nørremark for data management, to Anders Helles Carlsen and Maria Speed for statistical support, and to Torben Schmidt Kjeldsen for graphical design of the survey experiment. All of those acknowledged are affiliated with the Central Denmark Region.

## Data sharing

The data cannot be shared as the participants have not consented to data sharing.

