## Supplementary material for "Receiving information on machine learning-based clinical decision support systems in psychiatric services may increase patient trust in these systems: A randomised survey experiment"

### TABLE OF CONTENT

**PART 1: ENGLISH VERSIONS OF THE QUESTIONNAIRE AND THE ELECTRONIC INFORMATION PAMPHLETS**

1.1. Supplementary Table 1: Baseline questionnaire
1.2. Slides from "Intervention" and "Active control"
    1.2.1 Supplementary Table 2: Intervention
    1.2.2 Supplementary Table 3: Active control

**PART 2: SUPPLEMENTARY TABLES AND FIGURES**

2.1 Supplementary Table 4: Clinical characteristics of the 1,098 randomized participants
2.2 PCA analysis
    2.2.1 Supplementary Figure 1: Scree plot for trust items (positively worded outcome items)
    2.2.2 Supplementary Table 5: PCA component loadings for trust items (positively worded outcome items)
    2.2.3 Supplementary Figure 2: Scree plot for distrust items (negatively worded outcome items)
    2.2.2 Supplementary Table 6: PCA component loadings for distrust items (negatively worded outcome items)

2.3 Supplementary Table 7: Single items from the post-experimental questionnaire with t-tests between groups
2.4 Supplementary Table 8: Primary analysis stratified by age, sex, diagnostic category, educational level, work status

### PART 1: ENGLISH VERSIONS OF THE QUESTIONNAIRE AND THE ELECTRONIC INFORMATION PAMPHLETS

#### 1.1 Supplementary Table 1: Baseline questionnaire

| | | |
|---|---|---|
| | **Question/Statement** | **Options separated by commas** |
| **1** | What is your highest attained educational level? | Primary and lower secondary school, High school/Upper secondary school, Vocational school, Short-cycle higher education (less than 3 years of study),<br>Medium-cycle higher education (3-4 years of study),<br>Long-cycle higher education (more than 4 years of study), Other |
| **2** | What describes your current employment status the best? | Student (full time), Student (part time), Full-time employed, Part-time employed, Subsidized employment, Sick leave, Disability pension, Vocational rehabilitation, Unemployed, Retired, |
| **3** | How many adults are living in your household, besides yourself? | 0,1,2,3,4,5 or more |
| **4** | How many children are living in your household? | 0,1,2,3,4,5 or more, |
| **5** | Generally speaking, do you think that most people can be trusted or that you cannot be too careful in dealing with other people? | Likert scale 0-10<br>0: You cannot be to careful<br>10: Most people can be trusted |
| **6** | On a scale from 0 to 10, how much trust do you have in new technologies using computers/IT? | Likert scale 0-10<br>0: No trust at all<br>10: Full trust |
| **7** | On a scale from 0 to 10, how much trust do you have in each of the institutions listed below? | |
| **8** | The Danish parliament | Likert scale 0-10<br>0: No trust at all<br>10: Full trust |
| **9** | The judicial system | Likert scale 0-10<br>0: No trust at all<br>10: Full trust |
| **10** | The police | Likert scale 0-10<br>0: No trust at all<br>10: Full trust |
| **11** | European Union | Likert scale 0-10<br>0: No trust at all<br>10: Full trust |
| **12** | Healthcare services | Likert scale 0-10<br>0: No trust at all<br>10: Full trust |
| **13** | Psychiatric services in Central Denmark region | Likert scale 0-10<br>0: No trust at all<br>10: Full trust |
| **14** | Would you say that you understand how decisions are made regarding your treatment in psychiatry? | Likert scale 0-10<br>0: No understanding at all<br>10: Full understanding |
| **15** | Would you say you understand what machine learning models can potentially be used for in Psychiatry? | Likert scale 0-10<br>0: No understanding at all<br>10: Full understanding |

#### 1.2 Slides from “Active control” and “Intervention”

Every table cell was displayed as a single page in the electronic survey.

##### 1.2.1 Supplementary Table 2: Intervention

| |
|---|
| **Decision processes in the Psychiatric Services with support from machine-learning models**<br><br>On the following pages, you will find a brief explanation of how machine-learning models could support decision making in the Psychiatric Services of the Central Denmark Region. It is important that you read the entire material, as it could have an impact on the study. |

New technology makes it possible to use so-called machine-learning models to support decision making in the Psychiatric Services of the Central Denmark Region.

A new machine-learning model is developed by a computer being presented with material and examples that it can "learn from". Such material could be e.g. 10,000 electronic medical records from former patients (image 1). These records provide numerous examples of how a diagnosis is made. The computer (the machine) identifies patterns in these examples and "learns" to produce a qualified suggestion for a diagnosis based on the current patient record.
This process, in which a computer learns from examples, is called machine learning (image 2).

The resulting computer programme, which can prompt a suggestion for a diagnosis based on a patient record, is called a machine-learning model (image 3). The diagnosis suggested by the machine-learning model is presented to health personnel, who can take this information into consideration. These considerations also include the staff's own observations and results from various clinical examinations in addition to the suggestion made by the machine-learning model (images 4-5).

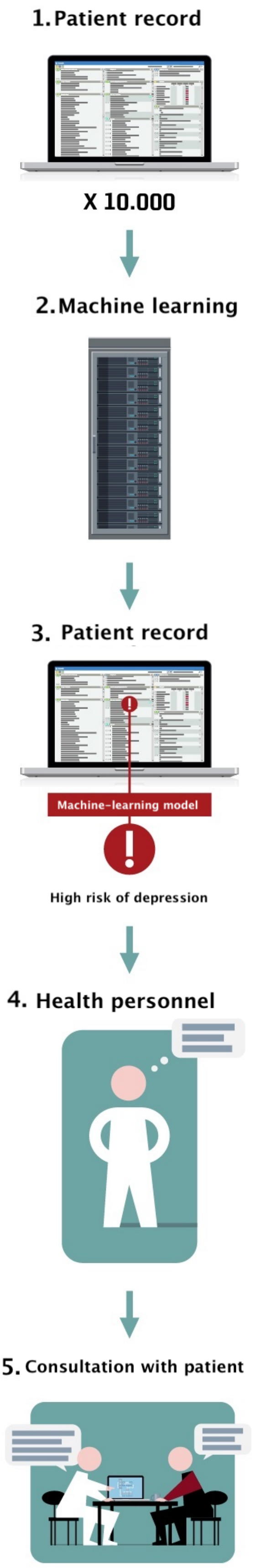

A decision-making process in a doctor-patient interaction in the Psychiatric Services could play out as follows:

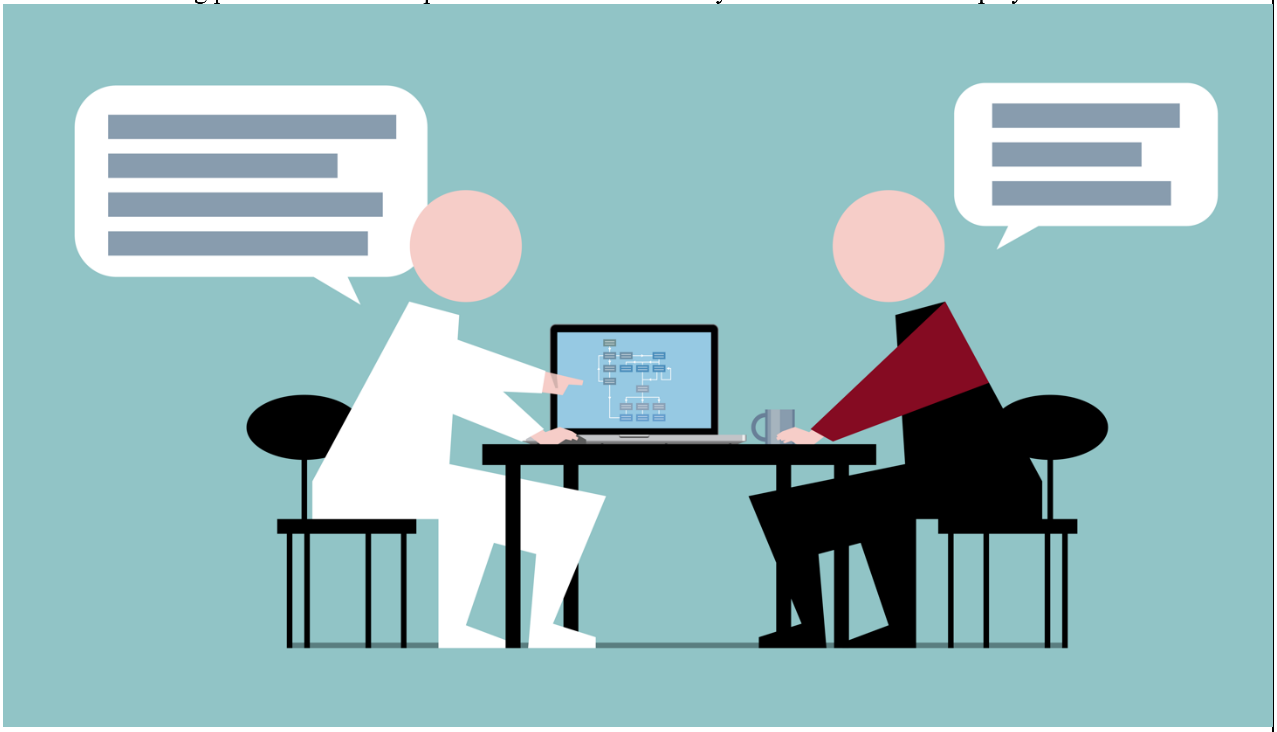

Doctor:
”Until now, we have been treating you for an anxiety disorder. But it can be difficult to differentiate between anxiety and depression. A computer programme in our system has analysed your patient record and suggested that you might have a depression. I have thought about it, and I am inclined to agree. Therefore, I would like to ask you some questions about symptoms of depression. Would it be OK with you if we do this now?”

Patient:
”I have also considered that myself. I am ready to answer the questions.”

The computer programme (machine-learning model) that helped the doctor in the example above focuses on diagnostics (detection of depression). However, machine-learning models may also offer guidance in other areas of psychiatry, such as:

- Early detection of physical illness, e.g. diabetes or cardiovascular disease
- Choice of optimal treatment for the individual patient (also called "personalised medicine")
- Evaluation of exacerbation of depression or psychosis.

It is important to emphasise that machine-learning models can only be used for decision support when the machine-learning models have been shown to perform reliably.

It will always be the health professionals, not the machine-learning models, who will make the final decisions in psychiatry. Thus, there is a significant difference between decision support (based on the models) and the actual decision, which will always be made by health professionals.

The overall objective of using machine-learning models for decision support in psychiatry is to improve the precision and effectiveness of both the diagnosis and the treatment.

#### 1.2.2 Supplementary Table 3: Active control

| |
|---|
| **Decision processes in Psychiatry**<br><br>On the following pages, you will find a brief explanation of how machine-learning models could support decision making in Psychiatric Services of the Central Denmark Region. It is important that you read the entire material, as it could have an impact on the investigation. |
| In the Psychiatric Services, many decisions are made regarding the treatment of the individual patient. These decisions are based on a broad range of data (investigations, consultations, patient preferences, etc.). Therefore, these decisions are complex.<br><br>The staff's decision-making process is often guided by information entered in the electronic patient record in connection with the patient's prior interactions with the Psychiatric Services.<br>The illustration below shows some examples of information in the patient record.<br><br>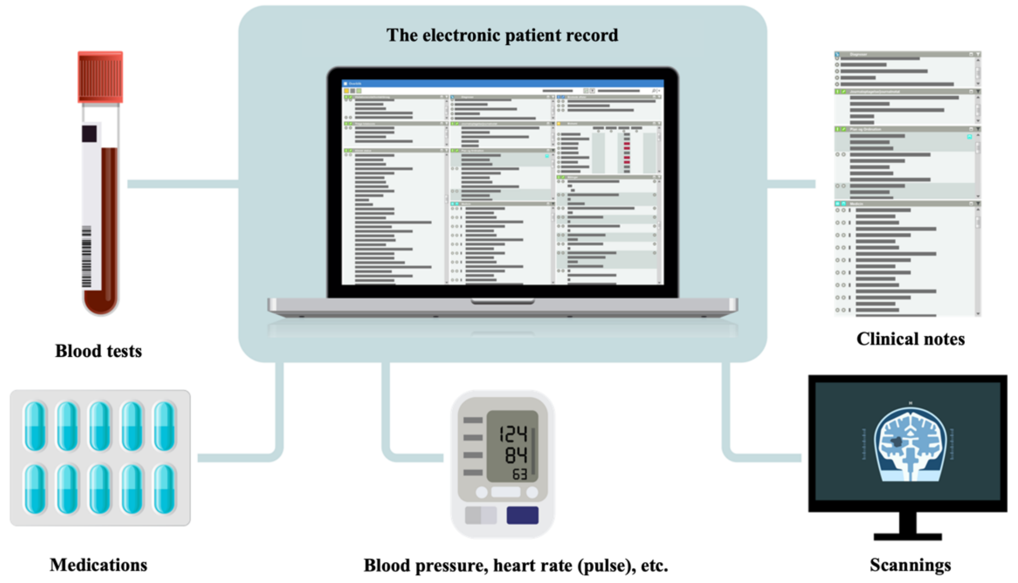 |
| Before a consultation with a patient, the staff prepares for the consultation based on entries in the record about the patient's prior interactions with the Psychiatric Services. A decision-making process in a doctor-patient interaction in |

the Psychiatric Services could play out as follows:

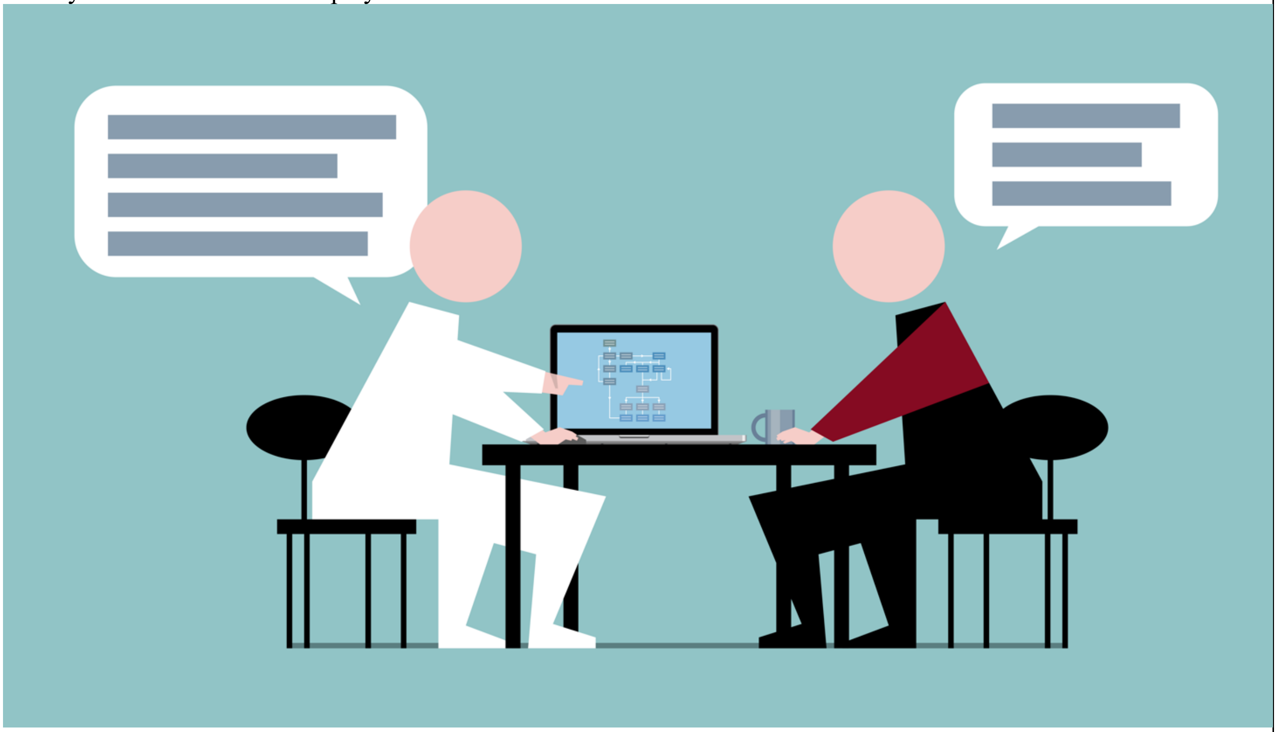

Doctor:
”Over the past weeks, we have been treating you for an anxiety disorder. But it can be difficult to differentiate between anxiety and depression. Based on our conversations and information from your patient record, I have found that I would like to ask you some more detailed questions about symptoms of depression. Would it be OK with you if we do this now?”

Patient:
”I have also considered that myself. I am ready to answer the questions.”

In the above example, a joint decision is made to investigate if the patient might have a depression. Other decisions could be:

- Ordering blood samples, e.g. infection rate or blood sugar
- Initiating/changing medications or psychotherapeutic treatment
- Completing a course of treatment

It will vary between patients what information forms the basis of individual decisions. A common feature of all decisions is that they aim to offer the best possible care for the individual patient.

PART 2: SUPPLEMENTARY TABLES AND FIGURES

### 2.1 Supplementary Table 4. Clinical characteristics of the 1,098 randomized participants

| Variable | Overall | Randomization group | | |
|---|---|---|---|---|
| | | **Blank Control** | **Active Control** | **Intervention** |
| | n=1,098 | n=356 | n=376 | n=366 |
| **Age,** median(IQR) | 34 (25) | 33 (25) | 35 (25) | 34 (25) |
| **Sex,** n (%) | | | | |
| Female | 779 (71%) | 245 (69%) | 272 (72%) | 262 (72%) |
| **Diagnosis*,** n (%) | | | | |
| Substance use disorder (F1) | 6 (1%) | <5 (<1%) | <5 (<1%) | <5 (<1%) |
| Psychotic disorders (F2) | 199 (18%) | 58 (16%) | 85 (23%) | 56 (15%) |
| Affective disorders (F3) | 603 (55%) | 207 (58%) | 199 (53%) | 197 (54%) |
| Neurotic disorders (F4) | 155 (14%) | 45 (13%) | 47 (13%) | 63 (17%) |
| Eating, sleeping and sexual disorders (F5) | 18 (2%) | <5 (<1%) | 7 (2%) | 7 (2%) |
| Personality disorders (F6) | 41 (4%) | 14 (3.9%) | 16 (4.3%) | 11 (3%) |
| Disorders of psychological development (F8) | 9 (1%) | <5 (<1%) | <5 (<1%) | <5 (<1%) |
| Child and adolescent disorders (F9) | 53 (5%) | 20 (6%) | 12 (3%) | 21 (6%) |
| Other | 14 (1%) | 5 (1%) | <5 (<1%) | 5 (1%) |
| **Total median contacts/year,** median (IQR) | 12 (12) | 11 (11) | 12 (12) | 13 (12) |
| **Days since first contact to Psychiatric Services,** median (IQR) | 1,293 (2753) | 1,179 (2542) | 1,565 (2902) | 1,260 (2836) |

*Most severe main diagnosis during the period with available data (2011-2023). (F*) indicates the ICD-10 chapter. Cell counts <5 are not specified due to risk of identification of individual patients.

### 2.2 Principal component analysis (PCA)

#### 2.2.1 Supplementary Figure 1: Scree plot for trust items (positively worded outcome items)

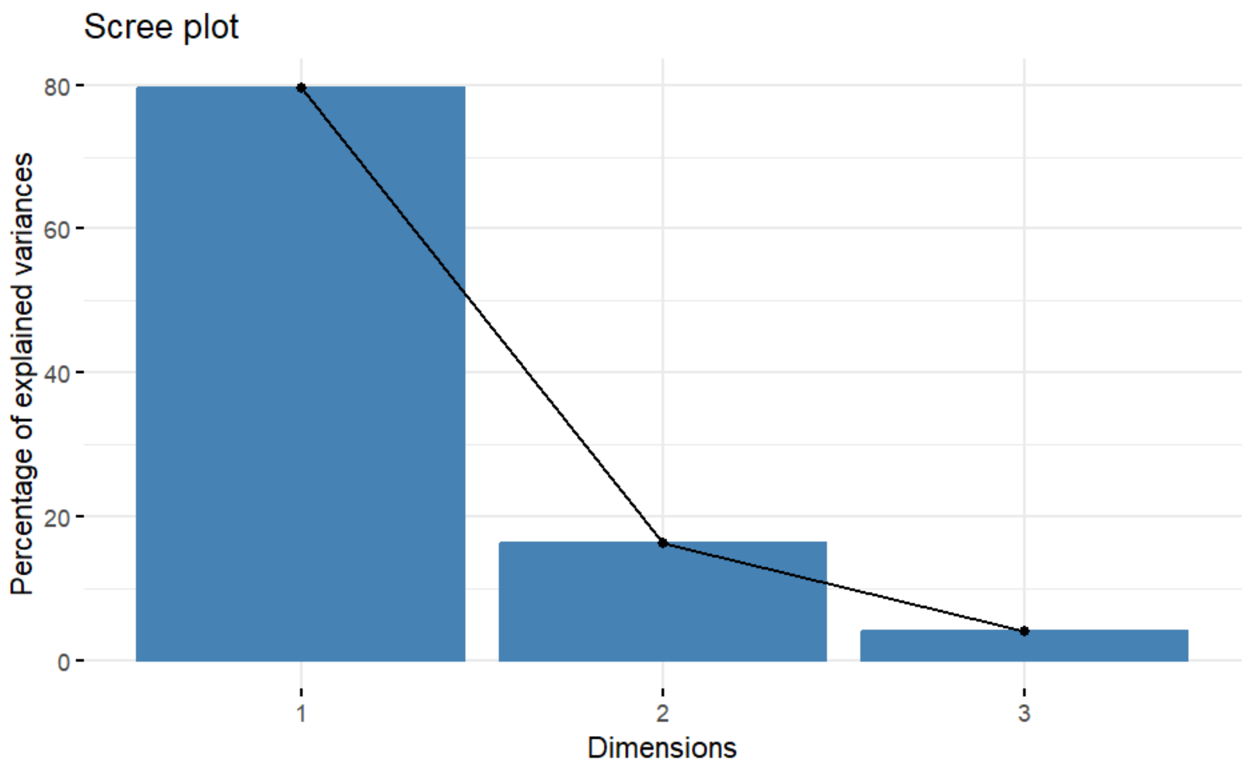

**2.2.2 Supplementary Table 5: PCA component loadings for trust items (positively worded outcome items)**

| | Component loadings | | |
|---|---|---|---|
| **Outcome Items (abbreviated*)** | PC1 | PC2 | PC3 |
| Item 1: I feel safe with ML models | 0.605 | -0.362 | 0.709 |
| Item 2: Trust in ML models, | 0.604 | -0.371 | -0.705 |
| Item 7: The advantages outweigh disadvantages | 0.518 | 0.855 | -0.006 |

*The full phrasing of the items and their scoring range are available in the methods section. PC: Principal component. ML: Machine learning.

**2.2.3 Supplementary Figure 2: Scree plot for distrust items (negatively worded outcome items)**

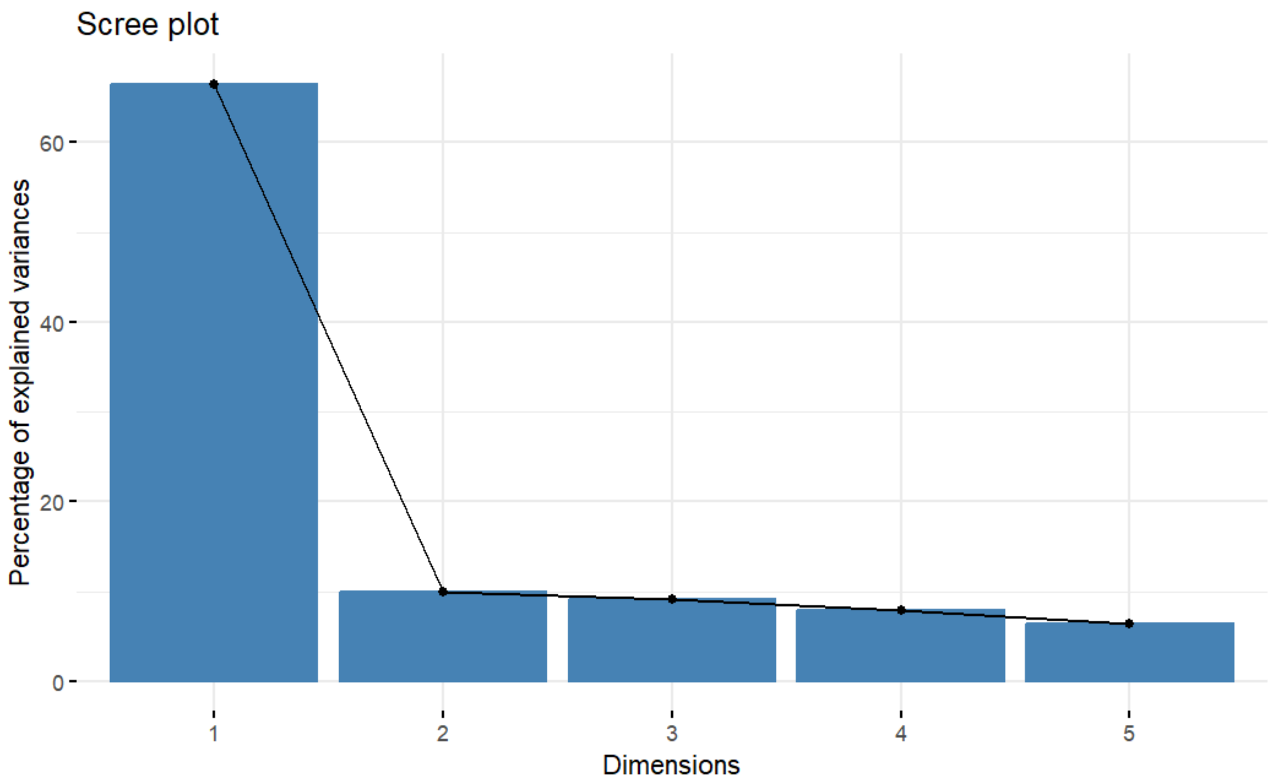

**2.2.2 Supplementary Table 6: PCA component loadings for distrust items (negatively worded outcome items)**

| | Component loadings | | | | |
|---|---|---|---|---|---|
| **Outcome items (abbreviated*)** | PC1 | PC2 | PC3 | PC4 | PC5 |
| Item 3: Malpractice due to ML | -0.452 | 0.182 | -0.268 | 0.808 | 0.195 |
| Item 4: Possibility to opt out of ML | -0.425 | -0.851 | -0.227 | -0.154 | 0.139 |
| Item 5: Dependency on ML | -0.474 | 0.131 | -0.061 | -0.106 | -0.862 |
| Item 6: Inequality in healthcare due to ML | -0.448 | 0.474 | -0.309 | -0.559 | 0.409 |
| Item 9: Errors in EHR | -0.436 | 0.011 | 0.882 | 0.002 | 0.179 |

*The full phrasing of the items and their scoring range are available in the methods section. PC: Principal component. ML: Machine learning.EHR: Electronic health record.

#### 2.3 Supplementary Table 7: Single items from the post-experimental questionnaire with t-tests between groups

| | Blank control vs. Active control | | Blank control vs. Intervention | | Active control vs. Intervention | |
|---|---|---|---|---|---|---|
| **Outcome items (abbreviated*)** | **Mean difference [CI]** | **p-value** | **Mean difference [CI]** | **p-value** | **Mean difference [CI]** | **p-value** |
| **Trust** | | | | | | |
| **I feel safe with ML models** | -0·088 [-0·517;0·342] | 0·69 | 0·503 [0·064;0·942] | 0·025 | 0·591 [0·159;1·02] | 0·0074 |
| **Trust in ML models** | 0·028 [-0·394;0·449] | 0·90 | 0·439 [0·008;0·869] | 0·046 | 0·467 [0·030;0·90] | 0·036 |
| **The advantages outweigh disadvantages of using ML models** | -0·002 [-0·412;0·408] | 0·99 | 0·466 [0·058;0·875] | 0·025 | 0·468 [0·053;0·884] | 0·027 |
| **Distrust** | | | | | | |
| **Dependency on ML models** | 0·264 [-0·212;0·740] | 0·28 | -0·114 [-0·598;0·370] | 0·64 | -0·378 [-0·869;0·103] | 0·12 |
| **Errors in EHR** | 0·014 [-0·413;0·440] | 0·95 | -0·368 [-0·809;0·073] | 0·10 | -0·382 [-0·820;0·057] | 0·088 |
| **Malpractice due to ML** | 0·054 [-0·385;0·492] | 0·81 | -0·778 [-1·22;0·333] | 0·00063 | -0·832 [-1·28;0·383] | 0·00030 |
| **Inequality in healthcare due to ML** | 0·075 [-0·396;0·546] | 0·75 | -0·359 [-0·833;0·115] | 0·14 | -0·434 [-0·902;0·034] | 0·069 |
| **Possibility to opt out ML** | -0·360 [-0·866;0·146] | 0·16 | -0·618 [-1·15;-0·086] | 0·023 | -0·258 [-0·782;0·265] | 0·33 |
| **Neutral** | | | | | | |
| **Explainability of a ML model** | 0·154 [-0·177;0·486] | 0·36 | 0·358 [0·017;0·698] | 0·039 | 0·203 [-0·159;0·566] | 0·27 |

*The full phrasing of the items and their scoring range are available in the methods section. ML: Machine learning. EHR: Electronic health record. Medians (IQR) for single items are shown in table 2.

### 2.4 Supplementary Table 8: Primary analysis for trust and distrust stratified by age, sex, diagnostic category, educational level, work status

| Trust | | | | | |
|---|---|---|---|---|---|
| Stratification variable | | Estimate | Standard Error | T-value | P-value |
| Age | | | | | |
| | Blank control vs Active control | -0.170 | 0.569 | -0.29 | 0.77 |
| | Blank control vs Intervention | 1.38 | 0.581 | 2.38 | 0.17 |
| | Active control vs intervention | 1.52 | 0.587 | 2.59 | 0.0097 |
| Sex* | | | | | |
| | Blank control vs Active control | -0.031 | 0.5628 | -0.056 | 0.95 |
| | Blank control vs Intervention | 1.44 | 0.5788 | 2.49 | 0.13 |
| | Active control vs intervention | 1.50 | 0.5835 | 2.58 | 0.010 |
| Diagnostic category | | | | | |
| | Blank control vs Active control | -0.092 | 0.572 | -0.161 | 0.87 |
| | Blank control vs Intervention | 1.42 | 0.584 | 2.42 | 0.016 |
| | Active control vs intervention | 1.51 | 0.591 | 2.56 | 0.011 |
| Educational level | | | | | |
| | Blank control vs Active control | -0.155 | 0.570 | -0.272 | 0.79 |
| | Blank control vs Intervention | 1.37 | 0.578 | 2.37 | 0.018 |
| | Active control vs intervention | 1.47 | 0.585 | 2.51 | 0.012 |
| Employment status | | | | | |
| | Blank control vs Active control | 0.088 | 0.572 | 0.154 | 0.88 |
| | Blank control vs Intervention | 1.49 | 0.580 | 2.57 | 0.010 |
| | Active control vs intervention | 1.44 | 0.589 | 2.45 | 0.015 |
| General trust | | | | | |
| | Blank control vs Active control | -0.195 | 0.558 | -0.35 | 0.73 |
| | Blank control vs Intervention | 1.41 | 0.570 | 2.472 | 0.014 |
| | Active control vs intervention | 1.61 | 0.574 | 2.82 | 0.005 |
| Baseline knowledge of machine learning | | | | | |
| | Blank control vs Active control | -0.009 | 0.530 | -0.016 | 0.99 |
| | Blank control vs Intervention | 1.72 | 0.555 | 3.099 | 0.002 |
| | Active control vs intervention | 1.78 | 0.557 | 3.188 | 0.002 |
| **Distrust** | | | | | |
| Stratification variable | | Estimate | Standard Error | T-value | P-value |
| Age | | | | | |
| | Blank control vs Active control | 0.110 | 0.970 | 0.113 | 0.91 |
| | Blank control vs Intervention | -2.06 | 0.970 | -2.126 | 0.034 |
| | Active control vs intervention | -2.27 | 0.985 | -2.306 | 0.021 |
| Sex* | | | | | |
| | Blank control vs Active control | -0.108 | 0.958 | -0.113 | 0.91 |

| | | | | | |
|---|---|---|---|---|---|
| | Blank control vs Intervention | -2.28 | 0.971 | 2.352 | 0.019 |
| | Active control vs intervention | -2.25 | 0.983 | -2.292 | 0.022 |
| Diagnostic category | | | | | |
| | Blank control vs Active control | -0.169 | 0.977 | -0.173 | 0.86 |
| | Blank control vs Intervention | -2.21 | 0.982 | -2.252 | 0.025 |
| | Active control vs intervention | -2.19 | 0.993 | -2.21 | 0.028 |
| Educational level | | | | | |
| | Blank control vs Active control | 0.217 | 0.967 | 0.224 | 0.82 |
| | Blank control vs Intervention | -2.17 | 0.971 | -2.231 | 0.026 |
| | Active control vs intervention | -2.26 | 0.984 | -2.30 | 0.022 |
| Employment status | | | | | |
| | Blank control vs Active control | -0.321 | 0.981 | -0.327 | 0.74 |
| | Blank control vs Intervention | -2.31 | 0.968 | -2.39 | 0.017 |
| | Active control vs intervention | -2.09 | 0.990 | -2.108 | 0.035 |
| General trust | | | | | |
| | Blank control vs Active control | 0.108 | 0.965 | 0.112 | 0.91 |
| | Blank control vs Intervention | -2.24 | 0.966 | -2.32 | 0.021 |
| | Active control vs intervention | -2.357 | 0.983 | -2.40 | 0.017 |
| Baseline knowledge of machine learning | | | | | |
| | Blank control vs Active control | 0.043 | 0.954 | -0.045 | 0.96 |
| | Blank control vs Intervention | -2.61 | 0.953 | -2.74 | 0.006 |
| | Active control vs intervention | -2.54 | 0.970 | -2.616 | 0.009 |

* Female is reference
